# Power and sample size considerations for test-negative design with bias correction: a case study on the world first malaria vaccine

**DOI:** 10.1101/2025.03.12.25323823

**Authors:** Yura K. Ko, Tobias Alfvén, Daisuke Yoneoka

**Affiliations:** Department of Microbiology, Tumor and Cell Biology (MTC), Karolinska Institutet, Sweden; Department of Virology, Tohoku University Graduate School of Medicine, Japan; Department of Global Public Health, Karolinska Institutet, Sweden; Sachs’ Children and Youth Hospital, Sweden; Center for Surveillance, Immunization, and Epidemiologic Research, National Institute of Infectious Diseases, Japan

**Keywords:** Test negative design, sample size, statistical power, vaccine effectiveness, malaria

## Abstract

**Background:** Test-negative design (TND) studies are increasingly common in evaluating vaccine effectiveness (VE) for various infectious diseases. TND studies are susceptible to bias due to disease outcome misclassification caused by imperfect test sensitivity and specificity. Several bias correction methods have been proposed. However, sample size or power considerations for TND studies incorporating bias correction for such misclassification have not yet been investigated.

**Methods:** Motivated by the world’s first malaria vaccine, we investigated how bias correction influences statistical power and sample size for VE estimation using Monte Carlo simulations. Under realistic assumptions about the proportion of vaccinated individuals in the general population, VE against clinical cases, and the probability of malaria diagnosis in unvaccinated individuals, we estimated the power to detect VE across different malaria vaccination statuses, with and without bias correction, at diagnostic test sensitivities of 60%, 80%, and 95%, and a specificity of 98%.

**Results:** The results demonstrated that using imperfect diagnostic tests reduces statistical power in both observed data-based VE and bias-corrected VE. The magnitude of power loss was substantially influenced by the sensitivity of the tests.

**Conclusions:** During the design phase of a TND study, researchers should conduct power calculations accounting for correcting the bias due to outcome misclassification. To achieve this, researchers need to collect comprehensive data, including the expected effect size of VE, the sensitivity and specificity of the diagnostic tests, the proportion of the vaccinated group, and the case ratio of the target disease.

## Main Background

Test-negative design (TND) studies are increasingly common in evaluating vaccine effectiveness (VE) for various infectious diseases^1^. Historically, the design has been applied to influenza vaccines^2^, and since the pandemic in 2020, it has been adopted in many countries, including those in Africa^3–5^, to estimate the VE of COVID-19 vaccines due to its efficiency. More recently, TND has been employed to evaluate novel vaccines for respiratory syncytial virus (RSV)^6^, started in 2023.

In a TND study, cases and controls are enrolled from the same location using the same clinical case definitions. This approach minimizes potential selection bias associated with health-seeking behavior compared to traditional case-control studies^1^. However, one of the limitations of TND is its vulnerability to misclassification of disease outcomes caused by the imperfect sensitivity and specificity of diagnostic tests^7^. Jackson et al. conducted a simulation study showing that influenza test misclassification led to greater VE underestimation in TND than in traditional cohort or case-control studies^7^.

To address this, Endo et al. proposed a bias correction method^8^ that can be applied using existing statistical software for logistic regression, the most common approach for estimating VE. They demonstrated that the correction method yields unbiased VE estimates, albeit with wider confidence intervals. Consequently, when less sensitive and/or specific tests, such as rapid diagnostic tests (RDTs), are used, a larger sample size is required to estimate unbiased VE with sufficient statistical power.

However, sample size or power considerations for TND studies incorporating bias correction have not yet been investigated.

In this study, we explore how the bias correction influences statistical power, using the malaria vaccine RTS,S/AS01, as a motivating example. There are two primary reasons for focusing on the malaria vaccine. First, RTS,S/AS01 is the world’s first malaria vaccine, recommended by the World Health Organization (WHO) in 2021 and prequalified in 2022. To date, no studies have evaluated the real-world VE of RTS,S/AS01 using TND. Second, in most clinical settings, the most common diagnostic tool for malaria is RDTs, which are highly specific but have relatively lower sensitivity^9,10^. This study aims to provide field epidemiologists, particularly those in African countries, with practical guidance for designing their first TND studies in such contexts. To facilitate this, we present a Shiny application that enables users to estimate VE by customizing their assumptions.

## Methods

We conducted Monte-Carlo simulations with the 500 iterations to estimate VE with and without bias correction and statistical power given different sample sizes (N = 5000, 6000, 7000, 8000, 9000, 10,000). All simulations and estimations were performed using Julia software (version 1.10.3). Reproducible code is available on GitHub (https://github.com/KoKYura/TND_power). Additionally, a Shiny application is available to calculate power by changing parameters and sample size (See the Supplementary file for detailed instructions on using the application).

### Simulation settings

We assumed the following data generation mechanism for the Monte-Carlo simulation.

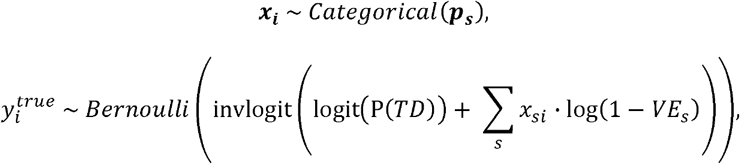

where 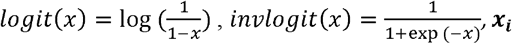 is the (one-hot) vector of binary indicator (i.e., 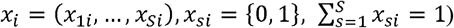 for vaccination status of individual, ***p***_*s*_ is the vector of proportions for each vaccination status among participants. The number of vaccination status can be unlimited (e.g, more than x days after vaccination, within x days after vaccination, and not vaccinated). However, as the categories being more detailed, the proportion for each status decreases, which in turn reduces statistical power. 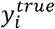 is the binary outcome of diagnosis using perfect tests, P(TD) represents the probability of being diagnosed with the target disease (TD) upon showing symptoms without vaccination, and *VE*_*s*_ ∈ [0,1]denotes the vaccine effectiveness against clinical illness for vaccination the probability of being diagnosed with the target disease (TD) upon showing symptoms without status *s*. As a key assumption in TND studies, we assumed that vaccines have no effect on the risk of non-target diseases (ND). Therefore, the proportion of each vaccination status among individuals with ND 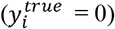 can be considered the same as the proportion of each vaccination status in the general population, which is obtainable data defined as ***p*** ^**′**^ _*s*._ Then, using the ratio of TD to ND for each vaccination status, *P*(*TD*_*s*_)/ *P*(*ND*_*s*_), we get the estimates of ***p*** ^**′**^ _*s*_ as (1+ *P*(*TD*_*s*_)/ *P*(*ND*_*s*_)).

Furthermore, assuming the TD and ND outcomes for 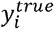 have (1-sensitivity) and (1-specificity) probabilities of being misclassified as false negatives or false positives, respectively, we derived 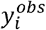, the binary outcome of observed diagnosis using imperfect tests.

The following assumptions were made to design a TND study for estimating the VE of the malaria vaccine RTS,S/AS01. The vaccine has a four-dose regimen with a three-dose primary series given at a minimum interval of four weeks between doses in children from five months of age, followed by a fourth dose 12–18 months after the third dose. Given the vaccination schedule and the relatively short duration of its effect^11^, the target population was defined as children aged two to five years. We assumed ***p*** ^**′**^ _*s*_ for this age range and VEs against clinical malaria at various intervals following each dose, as detailed in Table 1, based on previous clinical trial data^11,12^. We employed malaria RDT sensitivities of 60%, 80%, among individuals presenting with malaria-related symptoms without vaccine history *P*(*TD*))was and 95%, and a specificity of 98%, based on previous studies^9,10^,. The probability of malaria positivity assumed to be 30%, based on an ongoing cohort study (unpublished data)^13^ in Homa Bay County, Kenya.

**Table 1:** The assumption of general proportion among children aged 2–5 years old and VE against clinical malaria of each vaccination status.

| **Vaccination status** | **Proportion among**<br>**general population** | **VE against clinical**<br>**malaria** |
| --- | --- | --- |
| **Without dose 4** |  |  |
| more than 24 months after dose 3<br>or without dose 3 | 32.8% | 0%<br>(as reference) |
| up to 6 months after dose 3 | 2.6% | 50% |
| from 6 to 12 months after dose 3 | 2.6% | 30% |
| from 12 to 24 months after dose 3 | 4.5% | 10% |
| **With dose 4** |  |  |
| up to 6 months after dose 4 | 6.3% | 50% |
| from 6 to 12 months after dose 4 | 6.3% | 30% |
| more than 12 months after dose 4 | 44.9% | 10% |

### VE estimation with bias correction

We applied the multiple over-imputation method proposed by Endo et al.^8^ to obtain unbiased estimates of VE under the misclassification. Briefly, a parametric bootstrapping approach was employed, with the number of iterations set to 100, by flipping the observed test results at a certain probability. The VE for each vaccination status was then estimated, with 95% Confidence Intervals (CI) calculated based on 2.5^th^ and 97.5th percentiles of the estimates from the bootstrapped datasets. The detailed methodology is described elsewhere^8^.

### Power calculation and alpha error

Each statistical power for detecting true, observed, and bias-corrected VEs was defined as the proportion of datasets for which the 95% CI for the corresponding VE does not include Null value (i.e., 0).

Data sets were also simulated under the assumption of null VEs for all vaccination statuses to evaluate the probability of reporting a spurious VE where the vaccine has no effect. The alpha error was defined as the proportion of datasets under the null hypothesis for which the 95% CI for the corresponding VE includes 0.

## Results

The alpha error for each sample size, when bias correction was conducted for an imperfect diagnostic test with a sensitivities of 80%, 60%, and 95%, is presented in Table 2, Supplementary Table 1, and Supplementary Table 2, respectively. The results demonstrated that all simulations yielded an alpha error of less than 5%.

**Table 2:** The alpha error when the bias correction conducted for an imperfect test with a sensitivity of 80% and a specificity of 98% in each sample size for each vaccination status.

| N | -6 months,<br>Dose 3 | 6-12 months,<br>Dose 3 | 12-24 months,<br>Dose 3 | -6 months,<br>Dose 4 | 6-12 months,<br>Dose 4 | 12- months,<br>Dose 4 |
| --- | --- | --- | --- | --- | --- | --- |
| 5000 | 0.034 | 0.022 | 0.022 | 0.036 | 0.034 | 0.02 |
| 6000 | 0.02 | 0.018 | 0.028 | 0.028 | 0.028 | 0.016 |
| 7000 | 0.02 | 0.03 | 0.036 | 0.032 | 0.028 | 0.024 |
| 8000 | 0.042 | 0.028 | 0.018 | 0.028 | 0.03 | 0.024 |
| 9000 | 0.036 | 0.034 | 0.01 | 0.018 | 0.03 | 0.024 |
| 10,000 | 0.028 | 0.026 | 0.02 | 0.032 | 0.014 | 0.018 |

Figure 1, Supplementary Figure 1, and Supplementary Figure 2 show the estimated VEs with 95% coverage intervals across 500 simulations. In all vaccination status groups, the estimated VEs based on observed data were consistently underestimated. In contrast, the estimated VEs obtained using bias correction were unbiased but exhibited a wider range of estimates, which varied depending on the level of test sensitivity.

**Figure 1:**
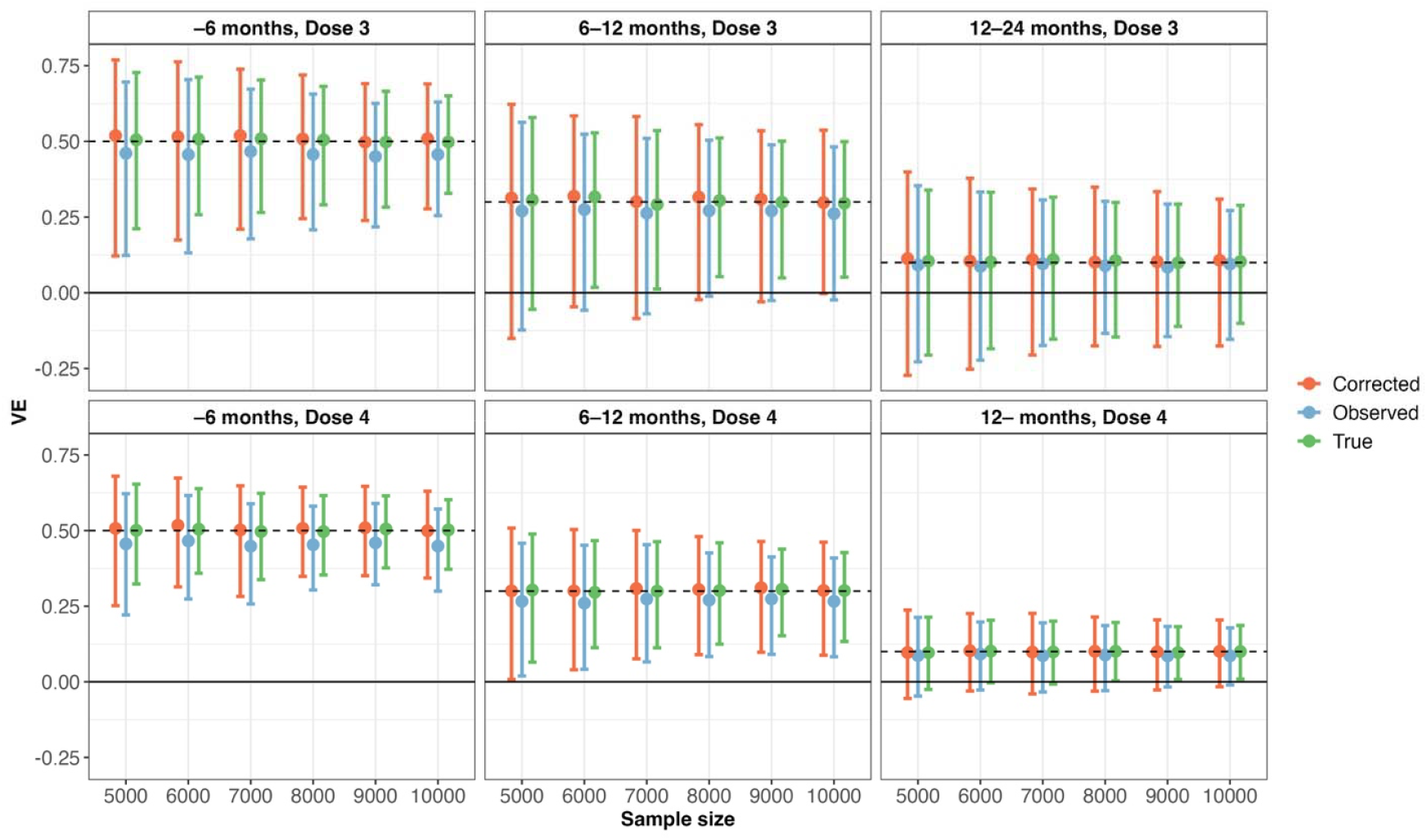
Estimated true, observed, and bias-corrected VEs for an imperfect test with a sensitivity of 80% and a specificity of 98%, presented with 95% coverage intervals across 500 simulations for each vaccination status group and sample size.

Figure 2 shows the estimated statistical power for each vaccination status and sample size. When estimating VE using bias correction for an imperfect test with a sensitivity of 60% or 80% and a specificity of 98%, the power was substantially reduced compared to the power estimated using a perfect test. The power was nearly equivalent to that of bias-uncorrected estimates for imperfect tests. For example, with a sensitivity of 80%, only three vaccination status groups (–6 months post Dose 3, –6 months post Dose 4, and 6–12 months post Dose 4) reached 80% power with a sample size of 10,000, while when using a perfect test, the power approached to 80% when the sample size was 6000 for these three groups. For highly sensitive tests (95% sensitivity), all three estimated VEs (true, observed, and bias-corrected) exhibited trivial differences in statistical power (the right panel of Figure 2).

**Figure 2:**
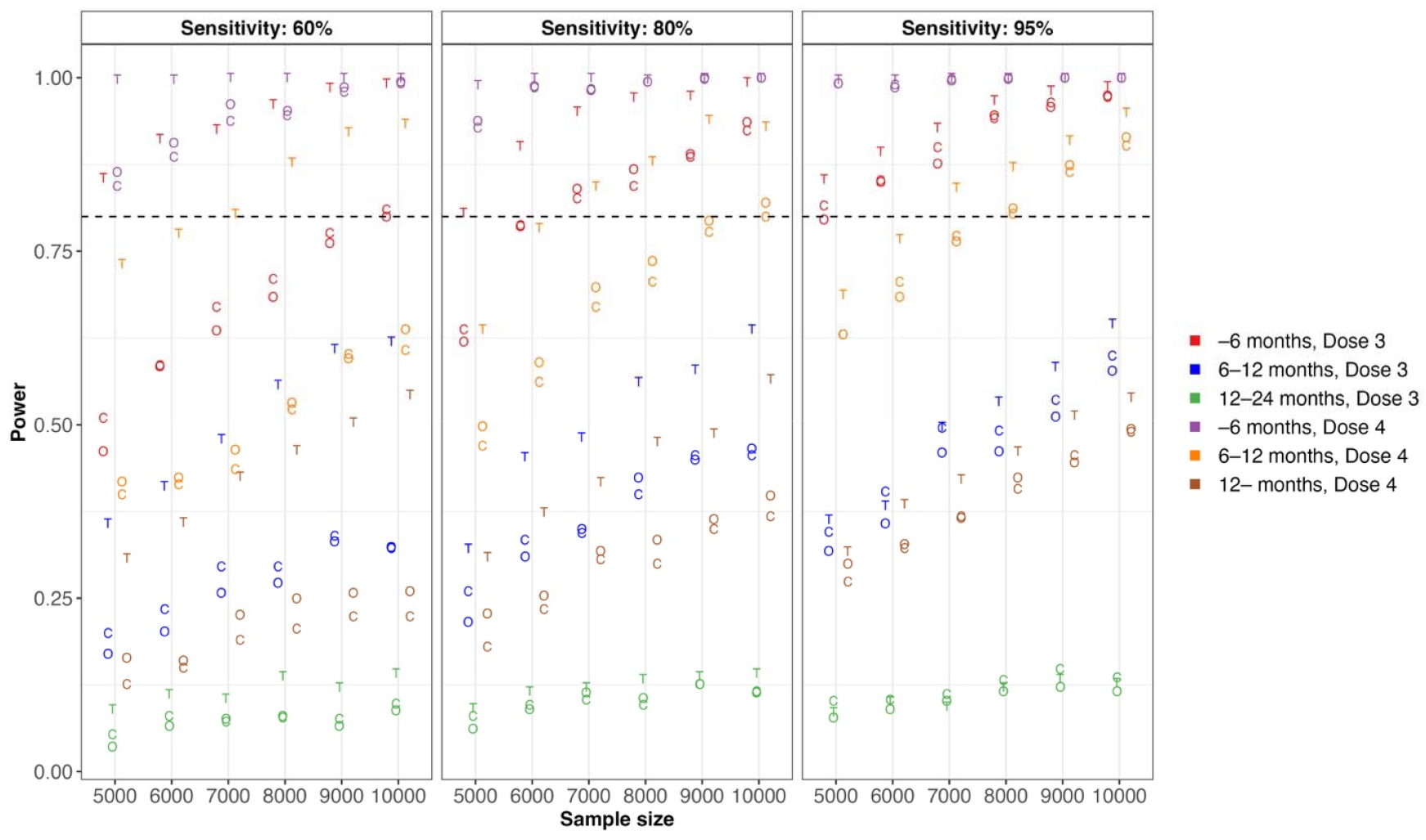
Estimated statistical power for true (shown as “T”), observed (shown as “O”), and bias-corrected (shown as “C”) VEs for an imperfect test with sensitivities of 60%, 80%, and 95%, and a specificity of 98%, across 500 simulations for each vaccination status group and sample size.

## Discussion

Our simulations demonstrated that using imperfect diagnostic tests reduces statistical power in both observed data-based VE and bias-corrected VE. The magnitude of power loss highly depends on the sensitivity of the tests. Given the limited research on power and sample size calculations for the TND^14^, our study provides valuable insights for assessing study feasibility, particularly for infectious diseases that are regularly diagnosed with less sensitive tests. This includes considerations of target populations, the number of study sites, and the duration of participant recruitment.

The bias in VE estimation due to the misclassification of outcomes in TND studies has been recognized for a long time^7,15,16^, and several bias correction methods have been proposed^8,17^. However, there are very few reports of VE estimates after correcting bias using these methods. For example, among the studies citing the work of Endo et al., only Amin et al. applied bias correction for sensitivity and specificity using the proposed method^18^, while Yoon et al. merely mentioned the bias as a study limitation^19^. Incorporating bias correction into the power calculation stage of research design could facilitate the wider adoption of bias-corrected VE estimation in future studies.

In this study, we used the world’s first malaria vaccine as a motivating example to simulate power for TND studies incorporating bias correction. Our simulation methods can also be applied to vaccines for other infectious diseases; however, several points should be addressed. First, our results indicated that higher VE corresponded to higher power. However, it is important to note that if VE is very high— such as above 90%, which is unlikely for malaria vaccines—the power may decline due to the limited number of vaccinated test-positive cases^14^. Second, we conducted simulations varying sensitivities (60%, 80%, 95%), while fixing the specificity at 98%. This decision was based on the fact that the specificity of RDTs is generally high, at 98% or above, including for malaria. Nevertheless, it should be noted that tests with lower specificity can have a greater impact on power^16^. Furthermore, although a closed-form solution exists for estimating bias-corrected VE proposed by Endo et al.^8^, we conducted Monte Carlo simulations; this was because the closed-form solution assumes a large-sample approximation, and the assumption may not always hold in the context of malaria, where sample size sparsity in certain vaccination statuses is expected.

This study has several limitations. First, we conducted a univariate analysis, incorporating vaccination status as the sole explanatory variable, rather than conducting a multivariate analysis. It should be noted that including additional variables as confounders would reduce the expected power^8^, depending on their effect size on the outcome and their (joint) distribution. Incorporating such assumptions into the simulation would require substantial prior information about the confounders, which is not feasible; therefore, we did not consider it in this study. Notably, the sample size calculation for evaluating the COVID-19 vaccine in TND, as published by the WHO, also assumes a univariate analysis^20^. Second, we considered a simplified scenario with constant vaccine coverage, no seasonal variation in TD risk, and, importantly, no change in infection prevention or risky behavior based on vaccination status. Although there is no clear evidence yet, if children vaccinated with the RTS,S/AS01 use mosquito nets less frequently in real-world settings, the effectiveness of the vaccine may be underestimated. In actual TND studies, it is important to conduct thorough interviews to assess these risky and preventive behaviors simultaneously. Third, we did not consider misclassification of vaccine status, which represents a potential source of bias in TND studies^21^. In the case of most childhood vaccines, vaccination records are maintained in the Mother-Child handbook. Ensuring that investigators verify these records, rather than relying solely on verbal responses, can minimize such misclassification.

In summary, we showed the simulated power for estimating bias-corrected VE when diagnostic tests have lower sensitivity using the malaria vaccine as a motivating example. During the design phase of a TND study, researchers should conduct power calculations accounting for correcting the bias due to outcome misclassification. To achieve this, researchers need to collect comprehensive data, including the expected effect size of VE, the sensitivity and specificity of the diagnostic tests, the proportion of the vaccinated group, and the case ratio of TD to ND. Such data can be obtained from pilot studies, published and/or unpublished data from the same region, and existing literature.

## Data Availability

All data produced in the present work are contained in the manuscript.

https://github.com/KoKYura/TND_power

## Ethics approval

Not applicable.

## Competing Interests

The authors declare no competing interests.

## Notes

### Competing Interest Statement

The authors have declared no competing interest.

### Funding Statement

This study did not receive any funding.

