## Supplementary file for "Power and sample size considerations for test-negative design with bias correction: a case study on the world first malaria vaccine"

### **Instructions for the shiny application**

To run the shiny application locally on your computer, follow these steps:

0. Install R and RStudio:

If you have not already installed R and RStudio, download and install them.

• **R**: <https://cran.r-project.org/>

• **RStudio**: <https://posit.co/download/rstudio-desktop/>

1. Download the shiny app files:

You can download all the necessary files from here (<https://github.com/KoKYura/TND_power>).

2. Install required R packages:

Open Rstudio and install the necessary packages by running the following commands in the R console.

Install.packages(“shiny”)

Install.packages(“shinydashboard”)

Install.packages(“DT”)

Install.packages(“JuliaCall”)

3. Run the application:

Open the “shiny/app.R” in Rstudio, and run the application either clicking the “Run App” button in Rstudio or running the following command in the R console:

shiny::runApp()

After running the command, you should see the following page.

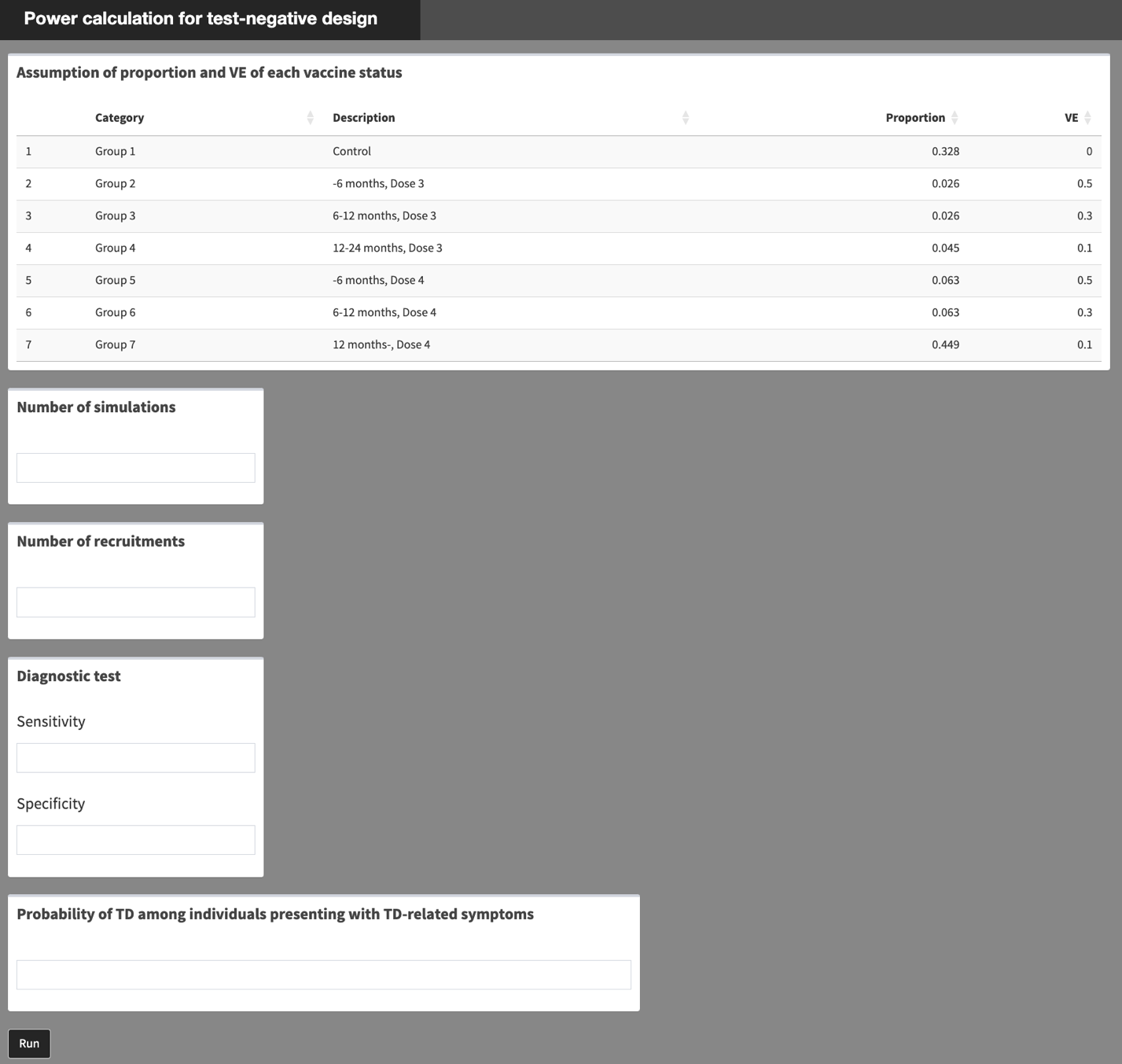

4. Change parameters and run simulations:

Based on the assumptions for your target diseases and study area, modify the following parameters:

- Proportion and vaccine effectiveness (VE) for each vaccination status
- Number of simulations
- Total number of recruitments
- Diagnostic test sensitivity and specificity
- Probability of being diagnosed as target disease (TD)

After setting all parameters, click “Run” to start the simulation. Once the simulation is complete, you will obtain the following plots.

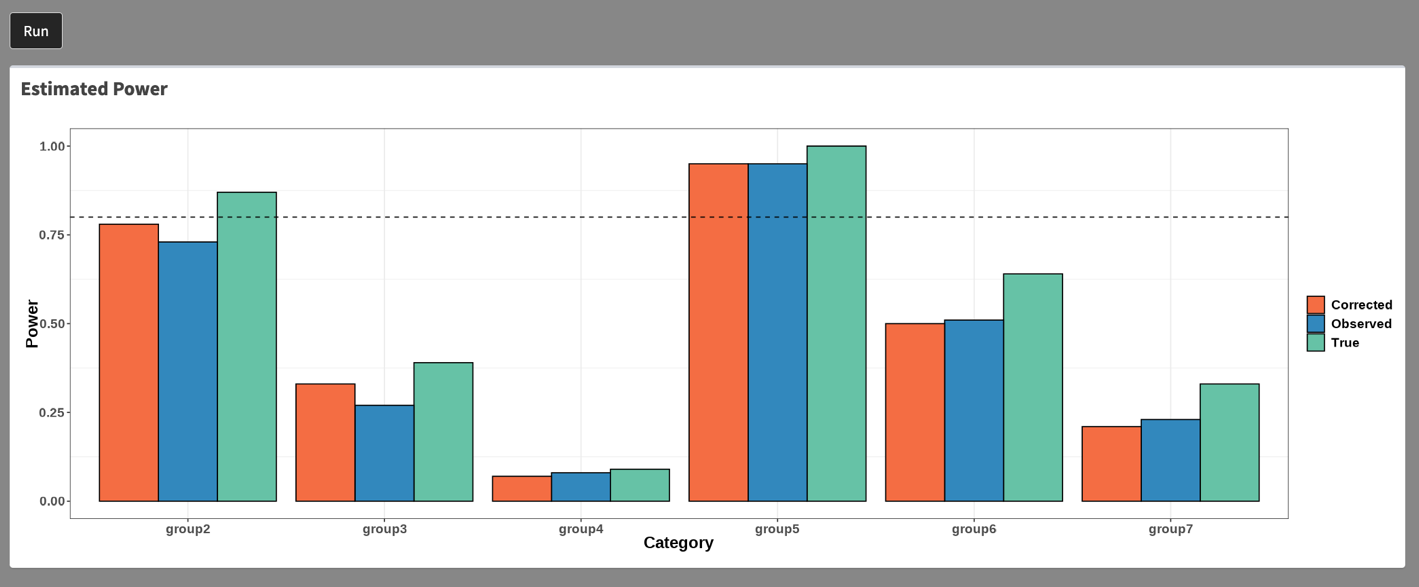

### **Supplementary Tables and Figures**

**Supplementary Table 1**: The alpha error when the bias correction cunducted for an imperfect test with a sensitivity of 60% and a specificity of 98% in each sample size for each vaccination status

| N | –6 months, Dose 3 | 6–12 months, Dose 3 | 12–24 months, Dose 3 | –6 months, Dose 4 | 6–12 months, Dose 4 | 12– months, Dose 4 |
| --- | --- | --- | --- | --- | --- | --- |
| 5000 | 0.032 | 0.026 | 0.024 | 0.026 | 0.032 | 0.02 |
| 6000 | 0.03 | 0.022 | 0.024 | 0.022 | 0.032 | 0.018 |
| 7000 | 0.022 | 0.026 | 0.026 | 0.024 | 0.034 | 0.014 |
| 8000 | 0.018 | 0.028 | 0.026 | 0.026 | 0.014 | 0.02 |
| 9000 | 0.036 | 0.024 | 0.016 | 0.03 | 0.014 | 0.026 |
| 10000 | 0.02 | 0.038 | 0.032 | 0.016 | 0.034 | 0.016 |

**Supplementary Table 2**: The alpha error when the bias correction cunducted for an imperfect test with a sensitivity of 95% and a specificity of 98% in each sample size for each vaccination status

| N | –6 months, Dose 3 | 6–12 months, Dose 3 | 12–24 months, Dose 3 | –6 months, Dose 4 | 6–12 months, Dose 4 | 12– months, Dose 4 |
| --- | --- | --- | --- | --- | --- | --- |
| 5000 | 0.034 | 0.03 | 0.038 | 0.028 | 0.016 | 0.01 |
| 6000 | 0.044 | 0.032 | 0.02 | 0.03 | 0.036 | 0.034 |
| 7000 | 0.048 | 0.03 | 0.036 | 0.036 | 0.022 | 0.024 |
| 8000 | 0.022 | 0.038 | 0.038 | 0.02 | 0.018 | 0.022 |
| 9000 | 0.024 | 0.028 | 0.022 | 0.02 | 0.024 | 0.022 |
| 10000 | 0.024 | 0.044 | 0.028 | 0.03 | 0.02 | 0.028 |

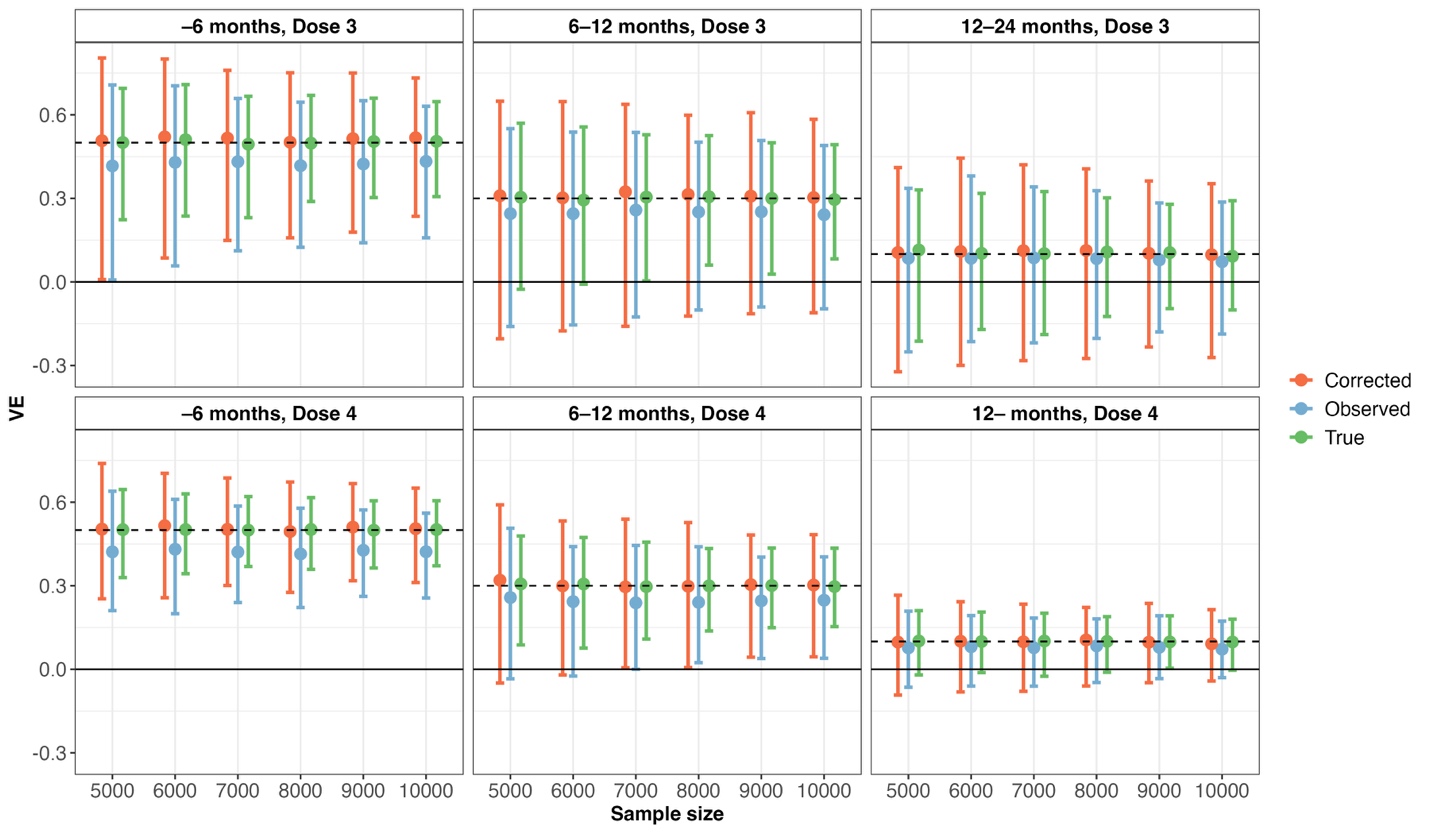

**Supplementary Figure 1**: Estimated true, observed, and bias-corrected VEs for an imperfect test with a sensitivity of 60% and a specificity of 98%, presented with 95% coverage intervals across 500 simulations for each vaccination status group and sample size.

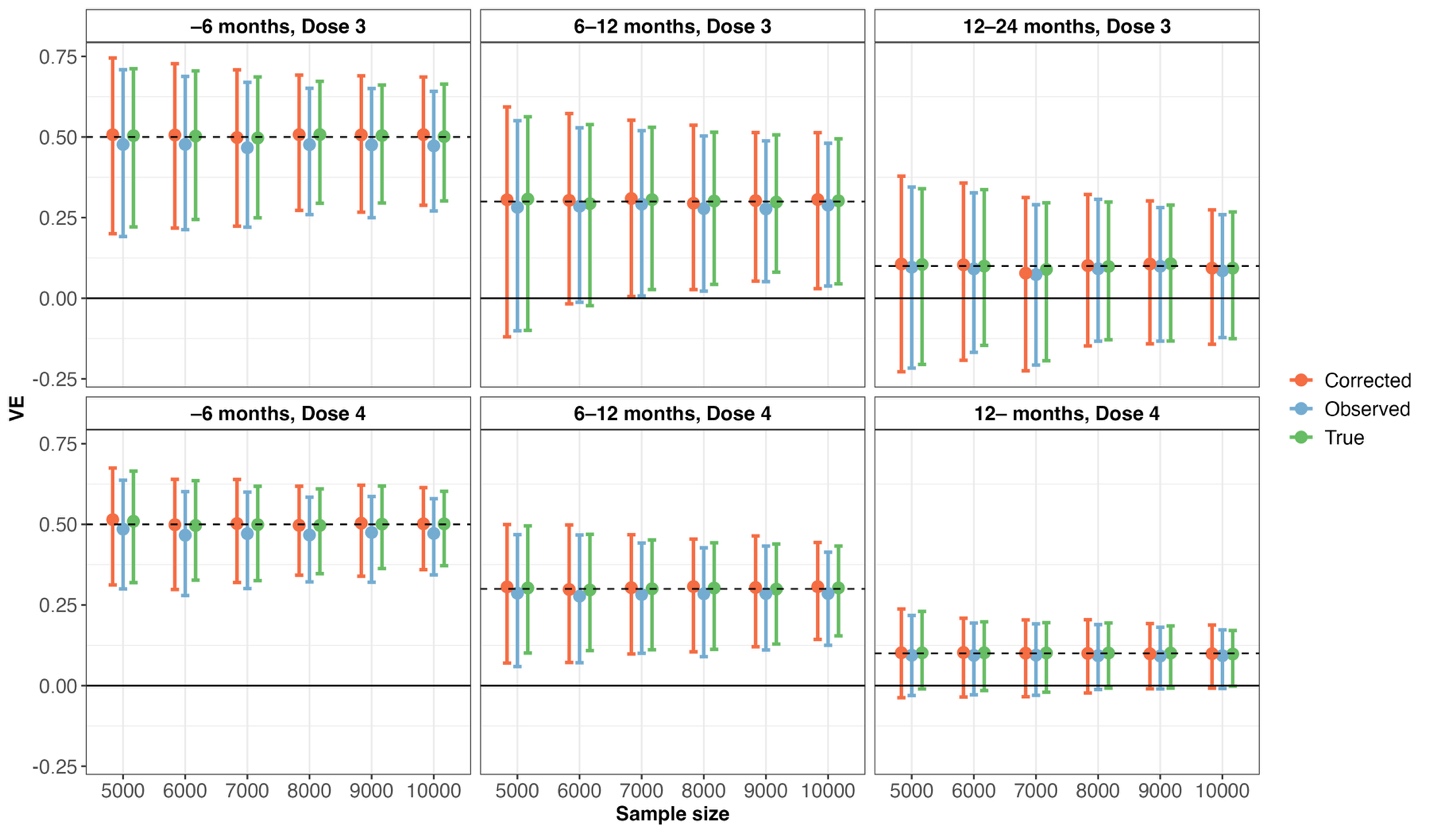

**Supplementary Figure 2**: Estimated true, observed, and bias-corrected VEs for an imperfect test with a sensitivity of 95% and a specificity of 98%, presented with 95% coverage intervals across 500 simulations for each vaccination status group and sample size.
